# Determinants of Binge Drinking Among Current Alcohol Drinkers: A Cross-Sectional Analysis of the 2013 Health and Welfare Survey

**DOI:** 10.64898/2026.11.03.26367174

**Authors:** Pongsin Pongsupatananon, Deanram Reangsan

## Abstract

This cross-sectional analytical study examined the prevalence and determinants of binge drinking (BD) among current alcohol drinkers (CAD) aged 20 years and older in Thailand (N = 52,847). Data from the Health and Welfare Surveys (HWS) were analyzed using chi-square tests and multiple logistic regression. In 2013, nearly 69% of the 5,445 current drinkers reported BD. Multivariable analyses indicated significantly higher odds of BD among men (AOR = 2.25, 95% CI: 1.90–2.66; *p* < 0.001), younger adults (AOR range: 1.50–1.90; *p* < 0.001), individuals with household housing expenses (AOR = 1.18, 95% CI: 1.01–1.39; *p* < 0.05), and those with secondary/vocational or tertiary education (AOR range: 1.35–1.36; *p* < 0.05). Smoking was also associated with elevated odds of BD (AOR = 1.42, 95% CI: 1.23–1.62; *p* < 0.001). In contrast, respondents in high-income groups had lower odds (AOR = 0.69, 95% CI: 0.54–0.87; *p* < 0.01), as did residents of non-metropolitan areas compared with Bangkok (AOR range: 0.44–0.59; *p* < 0.01). In conclusion, binge drinking affected over half of Thai drinkers and was strongly linked to demographic, socioeconomic, geographic, and behavioral factors. The frequent cooccurrence of alcohol use and smoking underscores the clustering of non-communicable disease risks, highlighting the need for integrated dual-cessation programs. While cross-sectional design and 2013 data limitations apply, these findings provide actionable evidence for public health agencies to implement targeted screening and intervention strategies for high-risk populations.

## Introduction

Alcohol consumption contributes to substantial health and social harms, including chronic diseases, traffic injuries, and interpersonal violence (WHO. 2024; Gandhi et al. 2024). Thailand has strengthened alcohol-control policies through the Thai Health Promotion Foundation Act (2001), which introduced a 2% surcharge on alcohol and tobacco products, followed by the Alcoholic Beverage Control Act B.E. 2551 (2008) and the National Alcohol Strategy (2011). These measures regulate alcohol availability and advertising, enforce drink-driving laws, restrict sales hours and locations, prohibit sales to intoxicated persons, and bar persons aged <20 years from pubs and nightclubs (Talek et al. 2024).

Despite extensive alcohol-control measures, per-capita alcohol consumption in Thailand increased from 7.4 to 8.2 liters between 2009 and 2013 (Global Health Observatory Data Repository, 2025). Alcohol use also imposes a substantial economic burden, estimated at 1.02% of Thailand’s gross domestic product (GDP). Premature mortality accounted for the largest proportion of these costs (95.45%), while healthcare expenditures constituted the second-largest component (2.7%) (Luangsinsiri et al. 2023). Binge drinking, characterized by episodes of heavy alcohol consumption, remains a particularly concerning pattern because of its acute health and social harms. (Assanangkornchai et al. 2010). To date, no study has specifically examined binge drinking among current drinkers using nationally representative Thai data. Focusing on this subgroup provides an important historical benchmark for understanding high-risk patterns that are often obscured in general population surveys. Accordingly, this study addresses two research questions:

1. What is the national prevalence of binge drinking among current alcohol drinkers in Thailand?
2. Which demographic, socioeconomic, geographic, and behavioral factors independently predict binge drinking within this subgroup?

## Research Objective

1. To estimate the prevalence of binge drinking among current alcohol drinkers aged 20 years and older.
2. To identify demographic, socioeconomic, geographic, and behavioral factors associated with binge drinking among current alcohol drinkers aged 20 years and older.

## Literature Review

Binge drinking is commonly defined as consuming alcohol to the point of intoxication, typically five or more standard drinks for men or four or more standard drinks for women on a single occasion (Courtney and Polich. 2009). This pattern of alcohol use is associated with both immediate and long-term harms, including alcohol-related problems and chronic liver conditions such as hepatitis and cirrhosis (Molina and Nelson. 2018). In Thailand, increasing alcohol consumption and the frequency of BD have contributed to the growing burden of non-communicable diseases (NCDs) (Wakabayashi et al. 2015).

Alcohol misuse is influenced by social and structural factors, including cultural norms, neighbourhood environments, and broader social determinants of health (WHO. 2024; Sudhinaraset et al. 2016). In Thailand, evidence highlights mixed socioeconomic patterns: regular BD is more frequent among individuals aged 11 and older in the lowest wealth quintile (Jankhotkaew et al. 2017), contrasting with trends among youth aged 12–15 (Prasartpornsirichoke et al. 2022). Furthermore, concurrent alcohol and tobacco use is disproportionately prevalent in lower-wealth groups (Intarut and Pukdeesamai. 2017), while older age (50–59 years) and smoking status are closely associated with CAD (Philalai et al. 2017).

Despite existing evidence, important gaps remain. Few studies (Nakhirunkanok. 2010; Jankhotkaew et al. 2017) have specifically investigated BD among CAD or examined its variation across demographic (younger and older adults), socioeconomic (household housing expense status, occupation), geographic (area of residence), and behavioral domains, including both health-risk and health-supportive practices. Addressing these gaps can inform more targeted alcohol-control policies for current drinkers. The 2013 Health and Welfare Surveys is uniquely valuable because it collected data on alcohol consumption and these factors, enabling their associations to be examined.

## Research Framework

Grounded in the social determinants of health (Solar and Irwin. 2010), the study examines binge drinking through four variable groups:

1. **Demographic Characteristics:** Age, sex, and marital status identify vulnerable sub-populations, such as younger adults and males, who historically exhibit higher risk-taking behaviors.
2. **Socioeconomic Status:** Monthly income and housing expense status reflect financial vulnerability and disposable income constraints (O’Donnell et al., 2008), while education and occupation serve as proxies for living standards.
3. **Geographic Factors:** Region and urbanization level account for regional disparities, cultural norms, alcohol availability, and localized policy environments.
4. **Behavioral Characteristics:** Smoking status, sugar-sweetened beverage intake, and health-promotion utilization capture the clustering of modifiable lifestyle risk factors.

**Figure 1.**
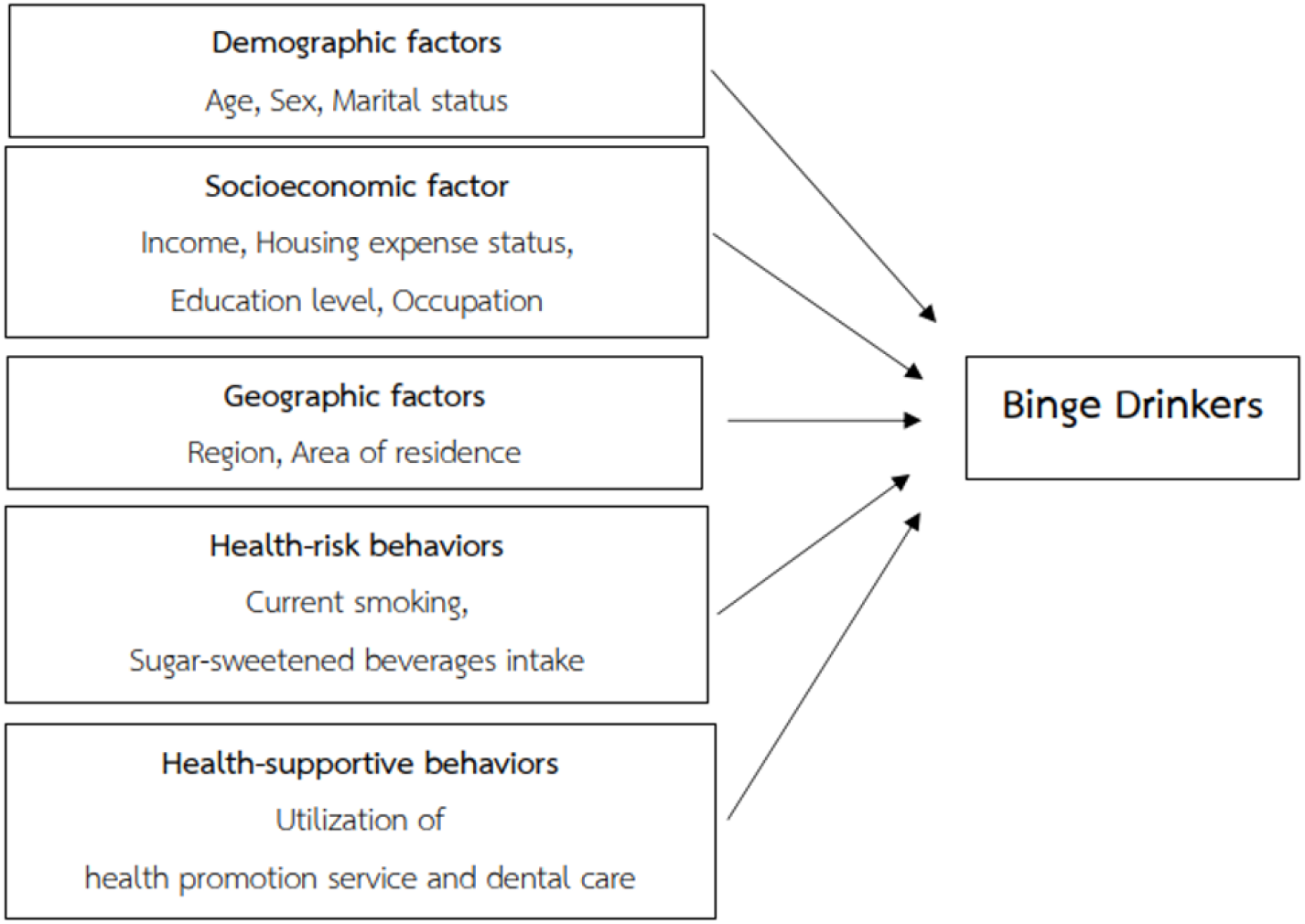
Research Framework.

## Research Hypotheses

### H1 (Prevalence)

BD accounts for the majority of the national prevalence among CAD aged ≥20 years in Thailand.

### H2a (Demographic)

Younger adults, males, and single marital status exhibit significantly higher odds of BD compared with older adults, females and married status.

### H2b (Socioeconomic)

Individuals with lower income, housing expense burdens, education levels and specific occupational categories show higher odds of BD.

### H2c (Geographic)

Residents of metropolitan areas (e.g., Bangkok) have higher odds of BD compared with those in non-metropolitan regions.

### H2d (Behavioral)

Smoking, sugar-sweetened beverage intake, lower utilization of health-promotion services and dental care are positively associated with BD.

## Research Methodology

### Research Design

This cross-sectional analytical study examined the national prevalence and associated factors of BD among CAD in Thailand. The analysis used secondary data from the Health and Welfare Survey (HWS), conducted by the National Statistical Office (NSO) of Thailand in March 2013.

### Population and Sample

This study used nationally representative data from the HWS. Although no a priori sample-size calculation was conducted, the large survey sample of 52,847 respondents provided adequate statistical power for population-level and stratified analyses. The study population included Thai adults aged ≥20 years in 2013, with non-drinkers excluded. The final complete-case analytic sample comprised 5,445 CAD.

**Figure 2.**
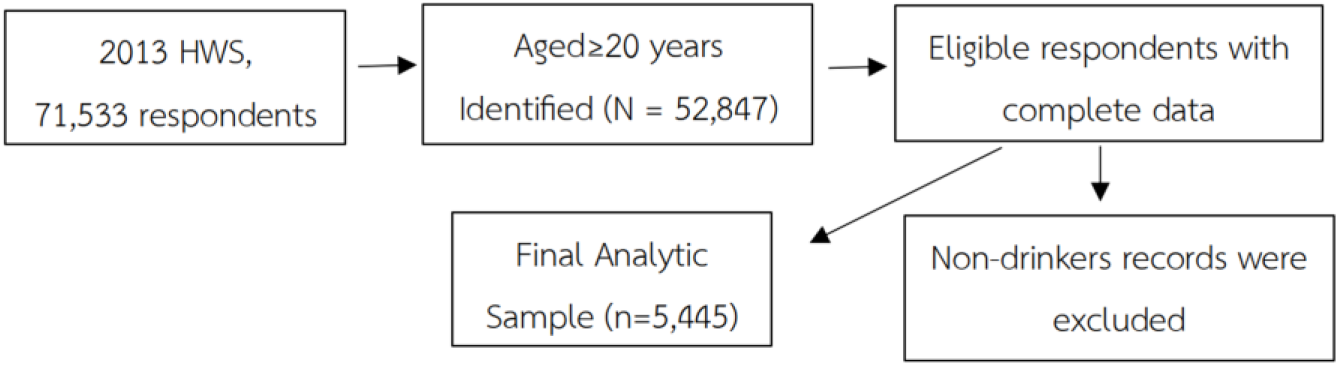
Sample Selection Flowchart.

### Research Instrument

Alcohol consumption was assessed using the HWS questionnaire. CAD were asked: “During the past 12 months, on days when you consumed alcohol, did you ever drink five or more glasses of liquor, beer, or wine, or nearly an entire jug of mixed spirits, on a single occasion?” Participants who answered “yes” were classified as binge drinkers. This threshold reflects drinking to intoxication in the Thai context (Lekskulchai and Rattanawibool. 2007) and is broadly consistent with international definitions of binge drinking (Courtney and Polich. 2009). Those who answered “no” were classified as non-binge drinkers (not shown in the Tables). For analysis, BD status was coded as a binary dependent variable (yes/no) and treated as the dependent outcome.

Independent variables were categorized into three domains: demographic, socioeconomic, and geographic characteristics. Demographic variables included age (20–39, 40–59, and ≥60 years), sex (male or female), and marital status (single/without a spouse or married). Socioeconomic characteristics included individual monthly cash income (≤7,000, 7,001– 20,000, and >20,000 Baht), categorized according to Wakabayashi et al. (2015), housing expense status (with housing expenses defined as households with ongoing rent, mortgage, and housing-related costs or without housing expenses defined as dwellings owned outright or occupied rent-free); educational attainment (primary education or lower, secondary or vocational education, and tertiary education or higher); and occupation. Income and housing expense status were included as direct indicators of living standards, consistent with O’Donnell et al. (2008). Occupation was classified using the standard occupational classification as economically inactive, skilled manual, skilled non-manual, or professional. Geographic characteristics included region (South, Northeast, North, Central excluding Bangkok, and Bangkok) and area of residence (municipal or nonmunicipal). Supplementary analyses evaluated health-risk behaviors, specifically current smoking (yes/no) and regular sugar-sweetened beverages (SSBs) intake (yes/no), and health-supportive behaviors, including the utilization of health-promotion services (e.g., vaccinations, health check-ups) and dental care services (yes/no), based on self-reported questionnaire responses.

### Data Collection

This study utilized secondary data from the 2013 Health and Welfare Survey (HWS), conducted by the National Statistical Office (NSO) of Thailand. The primary data collection was carried out by trained NSO field staff using structured household- and individual-level interviews. All study participants provided self-reported information. A two-stage stratified sampling design, incorporating survey weights, was used to generate nationally representative estimates. Primary sampling units consisted of residential blocks in municipal areas and villages in non-municipal areas. Within each selected block or village, households were randomly selected from official household registers and served as secondary sampling units.

### Data Analysis

Data were analysed using Stata version 18 (Stata Corp LLC, College Station, TX, USA). Survey weights accounted for unequal selection probabilities, nonresponse, and post-stratification calibration to national population benchmarks. All analyses used complex survey procedures incorporating primary sampling units (residential blocks or villages), and stratification by area of residence. Analyses were conducted in two stages. First, chi-square tests assessed bivariate associations between BD and each independent variable, with statistical significance set at *p* < 0.05. Second, variables significant in the bivariate analyses (*p* < 0.05) were included simultaneously in multivariable logistic regression models. Model assumptions were verified using multicollinearity diagnostics, which confirmed an acceptable mean VIF of 1.21. Adjusted odds ratios (AORs) and 95% confidence intervals (CIs) were estimated to identify factors independently associated with binge drinking.

## Research Findings

### Prevalence of Binge Drinking Behaviors

Table 1 presents the bivariate associations between BD and demographic, socioeconomic, geographic, and health-related characteristics. In 2013, the overall prevalence of BD among CAD was 68.96%. BD was significantly more prevalent among adults aged 20–39 and 40–59 years than among those aged ≥60 years (*p* < 0.001). Men had higher prevalence than women (72.34% vs. 52.44%, *p* < 0.001). Socioeconomic disparities were also evident. BD was more prevalent among respondents with middle or high incomes (*p* < 0.01), higher educational attainment, household with housing expenses (*p* < 0.001), and employment (*p* < 0.05), compared with their respective counterparts. Significant geographic variation was observed, with the highest prevalence in Bangkok (80.00%) and the lowest in the North (64.39%; *p* < 0.001). However, prevalence did not differ significantly between municipal and non-municipal areas. BD was also associated with other health-related behaviors. Current smokers had a markedly higher prevalence than non-smokers (74.54% vs. 64.52%, *p* < 0.001). Similarly, consumers of SSBs had a higher prevalence than non-consumers (70.28% vs. 64.44%, *p* < 0.001). In contrast, respondents who used health-promotion services had a lower prevalence of binge drinking than those who did not (61.49% vs. 69.71%, *p* < 0.05). Dental care utilization was not significantly associated with BD.

**Table 1.** Prevalence of binge drinking among current alcohol drinkers in 2013.

| **Determinants** | ***n*** | ***N*** | **%** | ***p*-value** |
|---|---|---|---|---|
| **Overall** | 3,755 | 5,445 | 68.96 |  |
| **Demographic characteristics** |  |  |  |  |
| Age: |  |  |  | <0.001 |
| 20–39 | 1,154 | 1,520 | 75.92 |  |
| 40–59 | 2,056 | 3,001 | 68.51 |  |
| ≥60 | 545 | 924 | 58.98 |  |
| Sex |  |  |  | <0.001 |
| Male | 3,271 | 4,522 | 72.34 |  |
| Female | 484 | 923 | 52.44 |  |
| Marital status |  |  |  | 0.428 |
| Single/without spouse | 1,094 | 1,568 | 69.77 |  |
| Married | 2,661 | 3,877 | 68.64 |  |
| **Socioeconomic characteristics** |  |  |  |  |
| Income (Baht) |  |  |  | 0.005 |
| ≤ 7,000 | 1,612 | 2,420 | 66.61 |  |
| 7,001–20,000 | 1,719 | 2,395 | 71.77 |  |
| >20,000 | 424 | 630 | 67.30 |  |
| Housing expense status |  |  |  | <0.001 |
| No housing expense | 2,651 | 3,958 | 66.98 |  |
| With housing expense | 1,104 | 1,487 | 74.24 |  |
| Education level |  |  |  | <0.001 |
| Primary or lower | 2,091 | 3,182 | 65.71 |  |
| Secondary/vocational | 1,048 | 1,401 | 74.80 |  |
| Tertiary or higher | 616 | 862 | 71.46 |  |
| Occupation |  |  |  | 0.012 |
| Economically Inactive | 319 | 506 | 63.04 |  |
| Skilled manual | 2,027 | 2,964 | 68.39 |  |
| Skilled non–manual | 882 | 1,233 | 71.53 |  |
| Professional | 527 | 742 | 71.02 |  |
| **Geographic characteristics** |  |  |  |  |
| Region |  |  |  | <0.001 |
| South | 420 | 631 | 66.56 |  |
| Northeast | 1,011 | 1,500 | 67.40 |  |
| North | 962 | 1,494 | 64.39 |  |
| Central (excluding Bangkok) | 1,150 | 1,555 | 73.95 |  |
| Bangkok | 212 | 265 | 80.00 |  |
| Area of residence |  |  |  | 0.741 |
| Non-municipal | 1,659 | 2,420 | 68.55 |  |
| Municipal | 2,096 | 3,025 | 69.29 |  |
| **Health-related behaviors** |  |  |  |  |
| Current smoker |  |  |  | <0.001 |
| yes | 1,798 | 2,412 | 74.54 |  |
| no | 1,957 | 3,033 | 64.52 |  |
| SSBs intake |  |  |  | <0.001 |
| yes | 2,963 | 4,216 | 70.28 |  |
| no | 792 | 1,229 | 64.44 |  |
| Health promotion service use |  |  |  | 0.049 |
| yes | 91 | 148 | 61.49 |  |
| no | 3,664 | 5,297 | 69.17 |  |
| Dental care use |  |  |  | 0.972 |
| yes | 372 | 540 | 68.89 |  |
| no | 3,383 | 4,905 | 68.97 |  |

**Notes:** *n* = number of BD within each subgroup; *N* = total number of respondents in each category. *p* < 0.05.

### Independent Predictors of BD Behaviors

Variables significantly associated with BD in the bivariate analyses were subsequently examined using univariable and multivariable logistic regression models. Univariable models estimated the crude association between each predictor and BD, whereas the multivariable model simultaneously adjusted for all selected variables to identify factors independently associated with BD among CAD (Table 2). Demographic characteristics revealed that compared to adults aged ≥60 years, those aged 20–39 and 40–59 years had significantly higher odds of BD (AOR range: 1.50–1.90; *p* < 0.001). Men had more than twice the adjusted odds of BD compared with women (AOR = 2.25, 95% CI: 1.90–2.66; *p* < 0.001).

**Table 2.** Crude and Adjusted Odds Ratios identifying independent determinants of BD among CAD.

| **Determinants** | **CORs** | **95% CIs** | ***p*-value** | **AORs** | **95% CIs** | ***p*-value** |
|---|---|---|---|---|---|---|
| **Demographic characteristics** |  |  |  |  |  |  |
| Age: |  |  |  |  |  |  |
| 20–39 | 2.19 | 1.80–2.67 | <0.001 | 1.90 | 1.52–2.39 | <0.001 |
| 40–59 | 1.51 | 1.28–1.79 | <0.001 | 1.50 | 1.25–1.80 | <0.001 |
| ≥60 | Ref. |  |  | Ref. |  |  |
| Sex |  |  |  |  |  |  |
| Male | 2.37 | 2.00–2.81 | <0.001 | 2.25 | 1.90–2.66 | <0.001 |
| Female | Ref. |  |  | Ref. |  |  |
| **Socioeconomic characteristics** |  |  |  |  |  |  |
| Income (Baht) |  |  |  |  |  |  |
| ≤ 7,000 | Ref. |  |  | Ref. |  |  |
| 7,001–20,000 | 1.27 | 1.11–1.47 | 0.001 | 0.87 | 0.75–1.03 | 0.101 |
| >20,000 | 1.03 | 0.81–1.31 | 0.797 | 0.69 | 0.54–0.87 | 0.002 |
| Housing expense status |  |  |  |  |  |  |
| No housing expense | Ref. |  |  | Ref. |  |  |
| With housing expense | 1.42 | 1.23–1.65 | <0.001 | 1.18 | 1.01–1.39 | 0.040 |
| Education level |  |  |  |  |  |  |
| Up to primary | Ref. |  |  | Ref. |  |  |
| Secondary/vocational | 1.54 | 1.36–1.77 | <0.001 | 1.36 | 1.16–1.59 | <0.001 |
| Tertiary or higher | 1.31 | 1.05–1.63 | 0.017 | 1.35 | 1.05–1.73 | 0.020 |
| Occupation |  |  |  |  |  |  |
| Economically Inactive | Ref. |  |  | Ref. |  |  |
| Skilled manual | 1.27 | 1.01–1.59 | 0.041 | 0.97 | 0.75–1.26 | 0.839 |
| Skilled non–manual | 1.47 | 1.16–1.87 | 0.002 | 1.02 | 0.79–1.32 | 0.888 |
| Professional | 1.44 | 1.07–1.92 | 0.014 | 0.99 | 0.73–1.36 | 0.960 |
| **Geographic characteristics** |  |  |  |  |  |  |
| Region |  |  |  |  |  |  |
| South | 0.50 | 0.30–0.82 | 0.007 | 0.44 | 0.27–0.72 | 0.001 |
| Northeast | 0.52 | 0.33–0.81 | 0.004 | 0.59 | 0.38–0.92 | 0.021 |
| North | 0.45 | 0.29–0.71 | 0.001 | 0.53 | 0.33–0.84 | 0.007 |
| Central (excluding Bangkok) | 0.71 | 0.48–1.13 | 0.145 | 0.72 | 0.46–1.14 | 0.164 |
| Bangkok | Ref. |  |  | Ref. |  |  |
| **Health-related behaviors** |  |  |  |  |  |  |
| Current smoker |  |  |  |  |  |  |
| yes | 1.61 | 1.41–1.83 | <0.001 | 1.42 | 1.23–1.62 | <0.001 |
| no | Ref. |  |  | Ref. |  |  |
| Daily SSBs intake |  |  |  |  |  |  |
| yes | 1.30 | 1.12–1.52 | 0.001 | 1.10 | 0.95–1.27 | 0.207 |
| no | Ref. |  |  | Ref. |  |  |
| Health promotion service use |  |  |  |  |  |  |
| yes | 0.71 | 0.49–1.03 | 0.070 | 0.83 | 0.57–1.22 | 0.345 |
| no | Ref. |  |  | Ref. |  |  |

**Notes:** (The Archer–Lemeshow goodness-of-fit test indicated adequate model fit, χ² (8) = 9.85, *p* = 0.275). CORs = crude odds ratios; AORs = adjusted odds ratios; CIs = confidence intervals; Ref. = reference category. The binary outcome was coded as 1 for binge drinkers and 0 for non-binge drinkers. *p* < 0.05.

Socioeconomic differences were also evident. Although crude BD rates were higher in middle-income groups than the lowest tier, multivariable adjustment showed high income was independently protective (AOR = 0.69, 95% CI: 0.54–0.87; *p* < 0.01). In contrast, individuals living in households with housing expenses had higher odds of BD than those without housing expenses (AOR = 1.18, 95% CI: 1.01–1.39; *p* < 0.05). Higher educational attainment was also associated with increased odds of BD compared with primary education or lower (AOR range: 1.35–1.36; *p* < 0.05). Occupation was not independently associated with BD after adjustment. Geographic differences also persisted. Residents of the North, Northeast, and South had lower odds of BD than Bangkok residents (AOR range: 0.4–0.59; *p* < 0.05), whereas the odds did not differ significantly between residents of Bangkok and the Central region. Health-related behaviors were independently associated with BD. Current smokers had higher odds of BD than non-smokers (AOR = 1.42, 95% CI: 1.23–1.62; *p* < 0.001).

## Discussion

This historical cross-sectional analytical study examined determinants of BD among Thai adults who were CAD in 2013. More than half of current drinkers reported BD, indicating a substantial burden of heavy and hazardous alcohol consumption and an elevated risk of intoxication and other alcohol-related harms. These findings underscore the need to strengthen and sustain alcohol-control measures to reduce harmful drinking. BD was independently associated with demographic, socioeconomic, geographic, and behavioral factors, thereby addressing the study objective.

Previous studies have identified peer influence as an important factor associated with alcohol consumption (Nakhirunkanok. 2010; Nonkhuntod and Yu. 2023). In occupational settings, alcohol use may be reinforced as a social norm through after-work gatherings and informal peer networks. This form of socialization may help explain the present finding that working-age adults had significantly higher odds of BD than older adults, including those who were retired.

Although international definitions of BD typically use a threshold of ≥4 drinks for women, the HWS measured BD only as consumption of ≥5 drinks. This may have underestimated BD among women and, consequently, exaggerated the observed sex disparity. Nevertheless, men had a higher prevalence of BD and more than twice the adjusted odds compared with women, consistent with previous research on BD (Nakhirunkanok. 2010) and among CAD in Thailand relative to those in other middle- and high-income countries (Sonthon. 2021, Chaiyasong et al. 2018).

Socioeconomic disparities in BD reflect a complex interplay of protective and risk factors. Among CAD, adjusted analyses indicate that markers of higher living standards, particularly high income and residence in households without housing expenses, are protective, consistent with patterns in the general Thai population (Jankhotkaew et al. 2017), likely due to reduced economic stress and greater access to health information. Conversely, higher education was linked to increased odds of BD among certain CAD, contrasting with trends reported in the general population (Wakabayashi et al. 2015). Although direct evidence from Southeast Asia is limited, this link may be driven by occupational networking and postwork socialization. In Thailand’s urban professional environments, alcohol often serves as a tool for corporate networking, team cohesion, and institutional loyalty. These habits transition from university subcultures into workplaces, where structured events normalize heavy drinking as part of career integration.

Regional disparities in alcohol consumption have been documented in Thailand (Chaiyasong and Thamarangsi. 2016). Consistent with this evidence, our findings indicate that BD is geographically patterned, with Bangkok metropolitan residents reporting the highest prevalence relative to those in other regions. This pattern may be related to a higher concentration of alcohol outlets, urban social norms surrounding drinking at social gatherings, and greater accessibility of alcoholic beverages in metropolitan than non-metropolitan areas.

Previous research suggests that alcohol consumption often co-occurs with other health-risk behaviors rather than occurring in isolation (Intarut and Pukdeesamai. 2017). Consistent with this evidence, our study found that BD was independently associated with current smoking. These findings highlight the clustering of alcohol use with other non-communicable disease risk factors.

## Study Strengths and Limitations

A key strength of this study is its use of nationally representative survey data and complex survey methods, allowing findings to be generalized to CAD in Thailand in 2013. The survey also included information on demographic, socioeconomic, behavioral, and health-related factors, enabling assessment of multiple correlates of binge drinking. However, the cross-sectional design precludes causal inference. Alcohol use was self-reported over a 12-month recall period and may be affected by recall and social desirability biases. In addition, the uniform five-drink threshold may underestimate BD among women, and comparable BD data were unavailable in other HWS waves, limiting assessment of temporal trends.

## Conclusions

Our findings indicate that more than half of CAD among Thai adults engage in BD, reflecting a substantial burden of heavy and hazardous alcohol consumption and an increased risk of intoxication and other alcohol-related harms. BD was also significantly associated with demographic, geographic, socioeconomic, and behavioral characteristics. Working-age adults, men, lower socioeconomic status, and residents of Bangkok should be prioritized because of their elevated risk. Targeted prevention and control measures for these populations may help mitigate subsequent harms. In addition, alcohol consumption commonly co-occurs with tobacco use, contributing to a clustering of modifiable risk factors for non-communicable diseases. Reducing binge drinking therefore requires targeted tobacco-alcohol dual cessation programs.

## Data Availability

All data produced in the present study are available upon reasonable request to the authors

https://www.nso.go.th/nsoweb/nso/survey_detail/96#gsc.tab=0

## Implications for Policy and Practice

1. Integrated dual-cessation programs: Align alcohol and tobacco control strategies to address their co-occurring, dual-risk behaviours simultaneously.
2. Targeted workplace and primary care Interventions: Expand routine screening and brief interventions in occupational and primary care settings, prioritizing working-age adults and male drinkers.
3. Urban availability regulations: Enforce zoning restrictions, outlet density limits, and operating hour controls in metropolitan centres such as Bangkok to reduce regionally concentrated binge drinking rates.

## Future Research Directions

Future studies should evaluate longitudinal trends in BD across Thailand to capture dynamic behavioral shifts over time. Additionally, research must clearly identify responsible agencies for specific policy measures, such as the Ministry of Public Health or local administrative organizations, to establish institutional accountability, streamline enforcement, and bridge the gap between policy formulation and practical implementation.

## Ethical Consideration

Data collection for the Health and Welfare Survey was conducted under the legal authority of the National Statistical Office in accordance with the Statistics Act B.E. 2550 (2007). The dataset used in this study was secondary in nature, contained no identifiable personal information, and all results were presented in aggregate form to ensure strict confidentiality of respondents.

## Acknowledgements

The authors would like to acknowledge the National Statistical Office of Thailand, Ministry of Digital Economy and Society (MDES) for providing the Health and Welfare Survey data, and are also grateful to the NSO’s staff for technical support.

